# The Every Mind Matters campaign in England: changes in mental health literacy over 30 months and associations between campaign awareness and outcomes

**DOI:** 10.1101/2022.11.08.22282079

**Authors:** Jane Sungmin Hahn, Kia-Chong Chua, Rebecca Jones, Claire Henderson

**Affiliations:** Division of Psychiatry, Faculty of Brain Sciences, University College London, London WC1E 6BT, UK; Health Service and Population Research Department, King’s College London, Institute of Psychiatry, Psychology, and Neuroscience, London SE5 8AF, UK

## Abstract

**Background:** The Every Mind Matters campaign and web resource launched in October 2019 by Public Health England aimed to equip adults to take action to improve their mental wellbeing by providing NHS-assured resources. The aim of this study is to investigate the effects on population level mental health literacy of Every Mind Matters over 30 months following campaign launch.

**Methods:** To observe changes in mental health literacy over time, we conducted regression analyses on a nationally representative, repeated cross-sectional dataset of nine survey waves from September 2019 to March 2022. We conducted an individual participant data meta-analysis with data from October 2019 to March 2021 to examine the association between campaign awareness and the outcomes, treating each survey wave as separate trials.

**Findings:** There were small improvements in knowledge of management of stress, depression, and anxiety, mental health vigilance, sleep literacy and psychological wellbeing self-efficacy from September 2019 to March 2020. By March 2022 there was a deterioration in all mental health literacy outcomes compared to September 2019, except for sleep literacy which was unchanged from baseline. Campaign awareness was positively associated with symptom management of depression and anxiety, help seeking self-efficacy, stigma related to mental disorders and mental health vigilance.

**Interpretation:** There is little evidence that the campaign improved mental health literacy in the general population beyond March 2020. Those who were aware of the campaign may have benefitted from its resources.

**Funding:** Public Health England, National institute for Health Research (NIHR) Policy Research Programme

**Research in context panel:** *Evidence before this study:* We used PubMed and Google Scholar to search for studies published between 2000 and 2022. We included papers that showed the relationship for mental health literacy (“mental health literacy, “stigma”, “help seeking”, “self-efficacy”), common mental health problems (“depression”, “anxiety”, “stress”, “sleep”, “low mood”, “common mental health problem”), mental health literacy interventions (“mental health literacy intervention”), and public health campaigns (“public health campaign*”, “anti-stigma campaign”, “mental health literacy campaign”). We also included studies discussing the mental health of the population because of COVID-19 (“population mental health”, “COVID-19”, “pandemic”, “wellbeing”), as lockdown started in the UK around six months into the campaign. Research into public health campaigns shows small-to-moderate improvements in mental health literacy, however, the campaign efforts have often been limited to stigma reduction. A meta-analysis found that Mental Health First Aid training led to small-to-moderate improvements in mental health first aid knowledge including recognition of mental health problems, beliefs about treatment, and attitudes related to stigma. Another meta-analysis found that web-based interventions could lead to improvements in mental health literacy if it included an active ingredient such as including evidence-based content or tailoring intervention to specific populations. However, these mental health literacy interventions often focussed on controlled settings rather than at a general population level. One study in Australia using national survey data found that public health campaigns focusing on a wider concept of mental health literacy than stigma improved beliefs about treatment and help seeking.

*Added value of this study:* This study adds value by evaluating a public health campaign at a population level and its effects based on a more comprehensive understanding of mental health literacy than has previously been operationalised. To do so, we used measures of mental health literacy developed to assess lay knowledge of daily life signs of depression, anxiety, and stress. Our study therefore has implications for stakeholders of the effectiveness of public health interventions, and whether these interventions can improve mental health literacy in the general population in a relatively short time span.

*Implications of all the available evidence:* The current evidence base indicates that sustained public health campaigns lead to small-to-moderate improvements in stigma related knowledge, attitudes and desire for social distance from people with mental health problems in the general population. However, we do not know whether all aspects of mental health literacy beyond help-seeking and stigma can be improved at population level through a campaign and web resource.

## Background

Mental Health Literacy (MHL) has been defined as 1) knowledge of how to obtain and maintain positive mental health, 2) knowledge of the symptoms and management of stress, anxiety, and low mood, 3) help seeking self-efficacy and the ability to promote one’s own mental health, and 4) stigma related to mental disorders. ^1^ MHL is associated with more effective mental health practices, ^2^ psychological wellbeing, ^3^ and outcomes related to common mental disorders^4^ in the general population. According to the Adult Psychiatric Morbidity Survey, in 2014, one out of six adults in England (ages 16-65) has a common mental disorder (CMD) such as depression or anxiety. ^5^ Since the last Adult Psychiatric Morbidity Survey, the COVID-19 pandemic has exacerbated multiple risks to mental health including financial insecurity, ^6^ social isolation, ^7^ bereavement^8, 9^ domestic abuse^10^ and occupational exposures to COVID. ^11, 12^ Therefore, promoting MHL at population level is an increasingly important public mental health goal.

Public Health England (PHE) developed a suite of mental health digital support resources and a promotional campaign, Every Mind Matters, which launched in October 2019. Its target was to help adults to take positive actions around their mental health. The digital resources comprise of the National Health Service (NHS)-assured content covering guidance on actions that people and the public can take to address the four most common sub-clinical mental health concerns: stress, anxiety low mood and sleep problems. Amongst these digital resources was the “Mind Plan”, which is an online questionnaire based on the Warwick-Edinburgh Mental Wellbeing Scale, ^13^ and it assesses the wellbeing of individuals and provide them with a tailored set of self-care actions to help them care for their mental health. By providing and encouraging the use of these digital resources PHE’s aim was to help prevent common mental health conditions from worsening and requiring NHS intervention.

Based on a pilot in the two Midlands regions of England in October 2018, PHE moved from a strategy that focussed on providing information about common mental health disorders to one that delivers and promotes evidence-based digital resources to facilitate self-care action for sub-clinical mental health problems for the national launch in October 2019. A second campaign under the Every Mind Matters banner across January/February 2020 encouraged people to talk openly about their mental health. A third campaign ran during April/May 2020 with a specific focus on promoting actions for people to take to care for their mental health during the first COVID-19 lockdown in response to a ministerial request. The fourth campaign in September/October 2020 changed direction to target parents rather than adults, with the aim of encouraging and supporting them to take action to look after the mental health of their children. This campaign strategy was complemented by advertising targeted directly at teenagers to help them look after their own mental health. Web analytics showed that between 7^th^ October 2019 and 28^th^ February 2021 the Mind Plan was completed 3,110,763 times.

Our approach to evaluation was based on the broad definition of MHL by Kutcher using the following outcomes: 1) symptom recognition and knowledge for symptom management of stress, anxiety, and depression, 2) mental health vigilance, 3) sleep-literacy 4) self-efficacy, and 5) stigma related to mental disorders. ^1^ Mental health vigilance fits into Kutcher’s definition of MHL as vigilance leads to the use of relevant knowledge in the maintenance of every day mental health and wellbeing. ^14^

Our objectives were therefore to 1) examine changes in knowledge of the symptoms and management of stress, anxiety, low mood, sleep literacy, help seeking self-efficacy and psychological wellbeing self-efficacy, stigma, and mental health vigilance among adults living in England from September 2019 to March 2021 and 2) compare these outcomes between those aware versus not aware of the campaign.

## Methods

### Data source

Members of the general population were recruited from a market research panel maintained by YouGov (https://yougov.co.uk/) to respond to an online survey prior to the launch of Every Mind Matters. This survey was repeated eight times after national launch. YouGov used quota sampling to create a sample of 20,000 respondents demographically representative of adults living in households in England.

### Measures

#### Exposures

We investigated change in MHL over nine survey waves (September 2019 n=3000, October 2019 n=2000, November 2019 n=2000, January 2020 n=2000, March 2020 n=3000, September 2020 n=2000, March 2021 n=2000, September 2021 n=2000, March 2022 n=2000).

Campaign awareness was assessed using the question “Do you remember seeing this ad recently?” Web, television, or radio advertisements were shown for the Oct 2019, Nov/Dec 2019, Jan 2020, Mar 2020, Sep 2020 and March 2021 campaign bursts. Participants responding that they had seen the content or something similar were categorised as being campaign aware. Those who answered that they had not seen the campaign or did not know they were categorised as campaign unaware. We did not include Sep 2019, Sep 2021, Mar 2022 as these waves did not measure campaign awareness.

#### Outcomes

Symptom recognition of stress, anxiety, and depression were measured by the Mental Health Literacy – Knowledge for Recognition (MHL-REC) scale. Symptom management of stress, depression and anxiety was measured by the Mental Health Literacy – Knowledge for Management (MHL-ACT) scale. ^14^ The MHL-REC consists of three identical sets of nine items and asks the participants to identify whether these experiences were a symptom of stress, depression, anxiety. Respondents could choose an answer for each condition or the option “none of these”. The scores range from 0 to 9 for each condition. The MHL-ACT asks whether each of seven actions could help with reducing each of depression, stress, and anxiety. The response format is the same as MHL-REC and scores range from 0-7 for each condition. Both the MHL-REC and MHL-ACT have been evaluated for construct validity. ^14^

Mental health vigilance was measured using the Mental Health Vigilance scale (MHL-VIG). ^14^ The MHL-VIG comprises 12 items on attitudes about maintaining mental health. The response options are ‘strongly disagree’, ‘disagree’, ‘neutral’, ‘agree’, and ‘strong agree’. Higher scores indicate a healthier attitude towards maintaining one’s mental health. MHL-VIG demonstrated good score reliability for discriminating a large range of individual differences. MHL-VIG data also demonstrated structural validity and convergent validity with constructs treated to mental health literacy such as symptom recognition, symptom management and mental health related stigma.

We measured sleep with the Sleep Beliefs Scale (SBS). ^15^ The scale includes 20 items divided into three factors: ‘sleep-incompatible behaviours’, ‘sleep-wake cycle behaviours’, and ‘thoughts and attitudes to sleep’. The total score ranges between 0 and 20. SBS shows good internal consistency of 0·71 (Cronbach’s alpha or CA). ^15^

We measured help-seeking self-efficacy using a subscale from the Mental Health Literacy Scale (MHLS) ^16^ and psychological wellbeing self-efficacy using a subscale of the Self-Rated Abilities for Health Practices Scale (SRAHPS). ^17^ The help-seeking subscale of the MHLS asks respondents to rate four statements of confidence in seeking help (e.g., “I am confident that I know where to seek information about in mental illness”). The response format is a Likert scale with 5 options (“Strongly disagree”, “Disagree”, “Neither agree or disagree”, “Agree”, and “Strongly Agree). MHLS shows good internal consistency of 0·87 (Cronbach’s alpha) and good test-retest reliability. ^16^ The psychological wellbeing subscale from SRAHPS consists of seven items which ask how well the respondent is able to do things that promote their mental health (e.g. “do[ing] things to make me feel good about myself” or “change things in my life to reduce stress”). Responses are “Not at all”, “A little”, “Somewhat”, “Mostly”, and “Completely”. The psychological wellbeing subscale of SRAHPS demonstrated high internal consistency (CA = 0·90) and moderate test-retest reliability (r=0·63). ^17^

We measured stigma related to mental disorders using intended behaviour items from the Reported and Intended Behaviour Scale (RIBS). ^18^ These items assess desire for social distance by enquiring whether the respondent would be willing to interact with someone with mental health problems in the future in four different contexts (living with, working, living nearby, and continuing a relationship). It uses a five-point Likert scale with ‘Agree strongly’, ‘Agree slightly’, ‘Neither agree or disagree’, ‘Disagree slightly’, ‘Disagree strongly’, as options. Scores from RIBS (ranging from 4 to 20) were reverse coded so that higher scores indicated less desire for social distance. YouGov also added an option for respondents to reply, “don’t know”, which was coded the same score as “Neither agree or disagree.” RIBS showed strong consensus validity as rated by service users, consumers, and international experts in stigma. It also demonstrated good internal consistency (CA = 0·85) and moderate to substantial test-retest reliability (r=0·75).

#### Demographic variables

We collected demographic data on age, gender, socioeconomic status and ethnicity (see Table 1). Socioeconomic status was categorised using the Market Research Society’s classification system into four groups, based on the occupation of the household’s chief income earner: AB = professional/ managerial occupations, C1 = other non-manual occupations, C2 = skilled manual occupations, and DE = semi-/unskilled manual occupations. Ethnicity was grouped into White, Mixed-White, Asian, Black, and Other based on UK census categories.

**Table 1:**
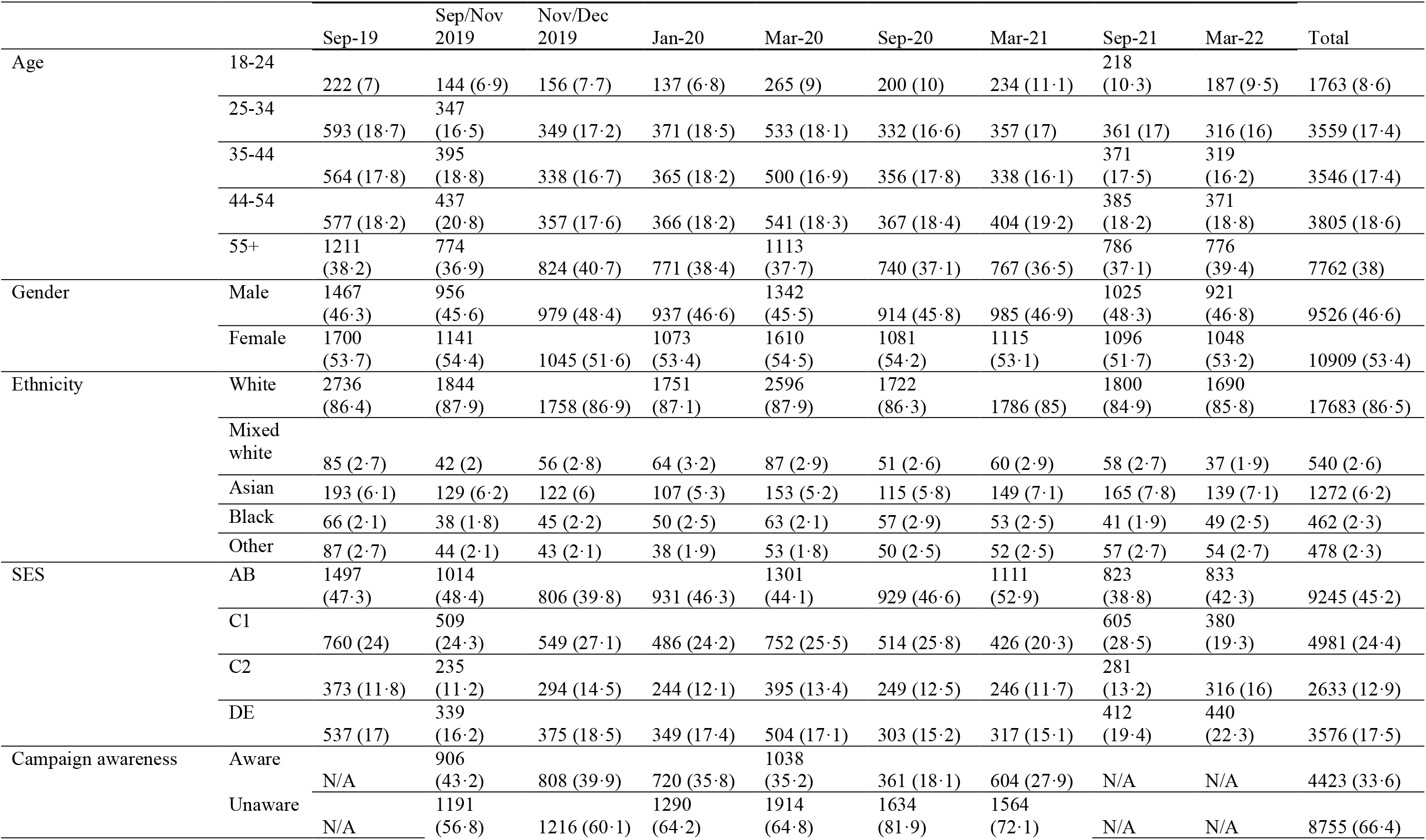
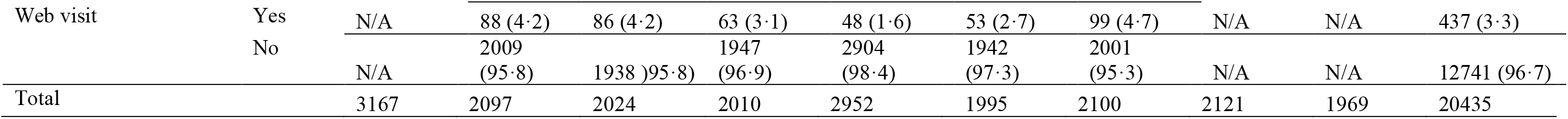
Demographic characteristics and campaign awareness over wave.

#### EMM web resource use

This was measured according to participants’ responses to the question “Before taking this survey, have you visited the Every Mind Matters website?” The participants were given a reminder about the contents including the online Mind Plan. Respondents answering “Yes” were categorised as having visited the website and not having visited if they answered “No” or “I don’t know”.

#### Statistical methods

To explore how MHL outcomes changed over time (objective 1), we used multivariable linear regression models with dummy variables representing seven survey waves of data-collection to assess the effect of time on 11 MHL outcomes. We controlled for demographic variables (age, sex, ethnicity and SES).

We expected the association between campaign awareness and MHL to differ amongst the survey waves for because: first, respondents in each wave were cross-sectional samples of the population, not the same cohort of individuals; second, measurement of campaign awareness in later waves included more targeted questions about Every Mind Matters materials on television radio, and the web; third, respondents at a later wave would have had greater average exposure to the campaign due to cumulative campaign activity. Therefore, to investigate the relationship between campaign awareness and MHL outcomes (objective 2) we used Independent Participant Data (IPD) meta-analysis to gain an overall picture from multiple waves of survey, accounting for between-study heterogeneity across survey sample and time points. We used a two-stage random effects model with restricted maximum likelihood (REML) estimation ^19^ for our IPD meta-analysis. We derived the 95% confidence intervals using the Hartung-Knapp-Sidik-Jonkman (HKSJ) approach, which produces more adequate error rates than other methods of estimation. ^20^ We treated the individual survey waves as different trials. Effect estimates were generated by using a linear regression model with awareness as the exposure and the MHL variables as the outcomes, adjusting for age, sex, ethnicity, and SES. We used τ2 to measure proportion of total variability due to between-study heterogeneity.

All analyses were conducted using Stata 16.

## Results

### Objective 1: Change over time

There was a total of 21,131 respondents over nine waves. The final analytical sample comprised 20,435 respondents. 696 respondents were excluded due to missing data on who answered “prefer not to say” on the ethnicity item. People aged over 55, those of white ethnicity and those in the professional/managerial occupation are the biggest groups within their respective demographic categories (Table 1)

All the results for change over time can be found in Table 2. There were small improvements in symptom management for stress (MHL-ACT stress) (estimated difference, d = 0·14; 95% CI: 0·02 to 0·25; p = 0·02) and depression (MHL-ACT depression) (d = 0·13; 95% CI 0·02 to 0·24; P = 0·02), and anxiety (MHL-ACT anxiety) (d = 0·21; 95% CI 0·1 to 0·32; P<0·001) in Mar 2020. Mental health vigilance (MHL-VIG) showed small improvements from Sep 2019 to Jan 2020 (d = 0·58; 95% CI 0·16 to 1; P = 0·007) and Mar 2020 (d = 0·59; 95% CI 0·22 to 0·97; P = 0·002). Sleep literacy (SBS) showed the most consistent and steady improvement from Sep 2019 to Nov/Dec 2019 (d = 0·32; 95% CI 0·1 to 0·53; P = 0·01) and to Sep 2020 (d = 0·44; 95% CI 0·22 to 0·66; P<.001). There was a small improvement for psychological wellbeing efficacy to Mar 2020 (d = 0·42; 95% CI 0·13 to 0·72; P = 0·005).

**Table 2:**
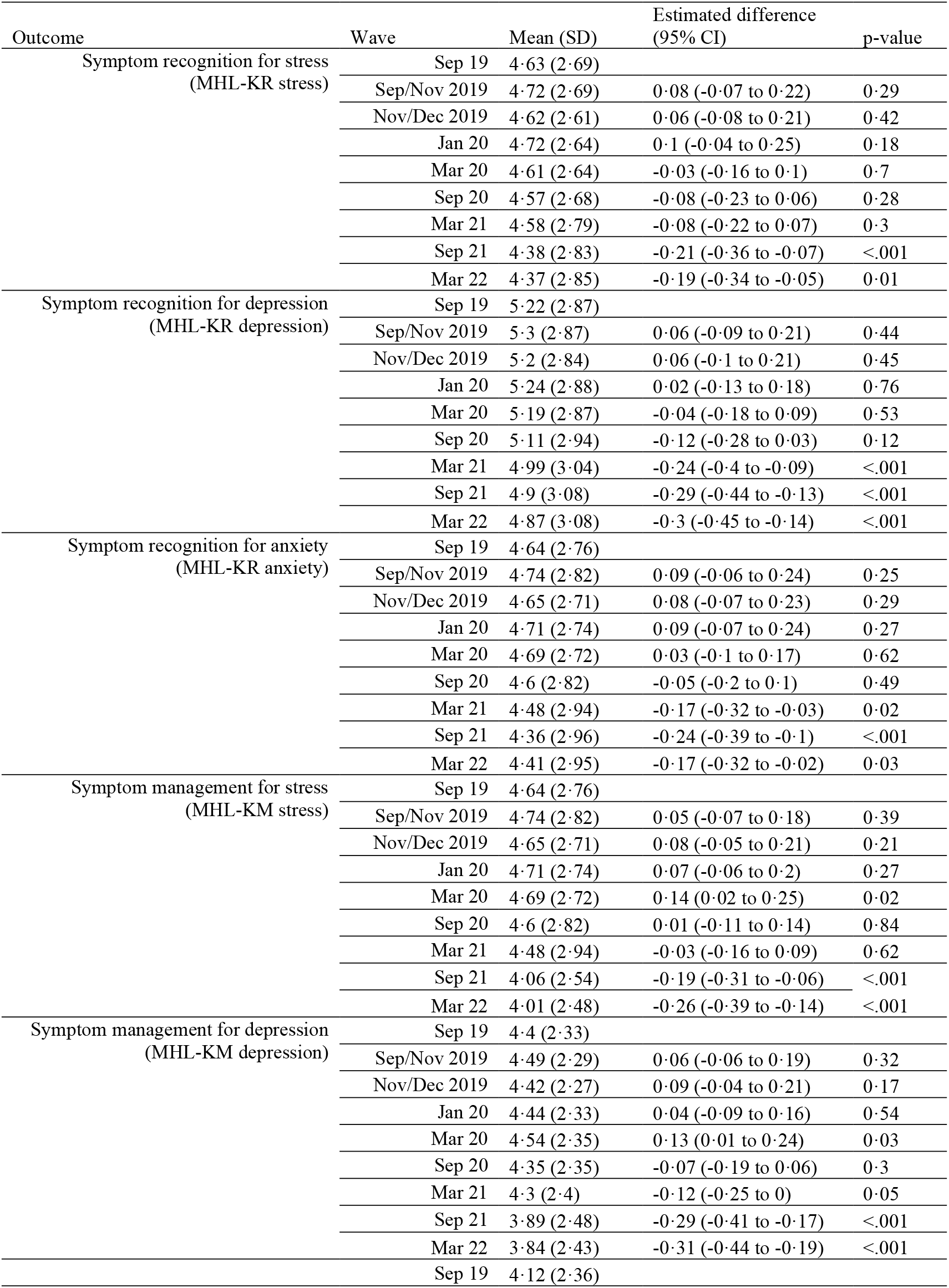
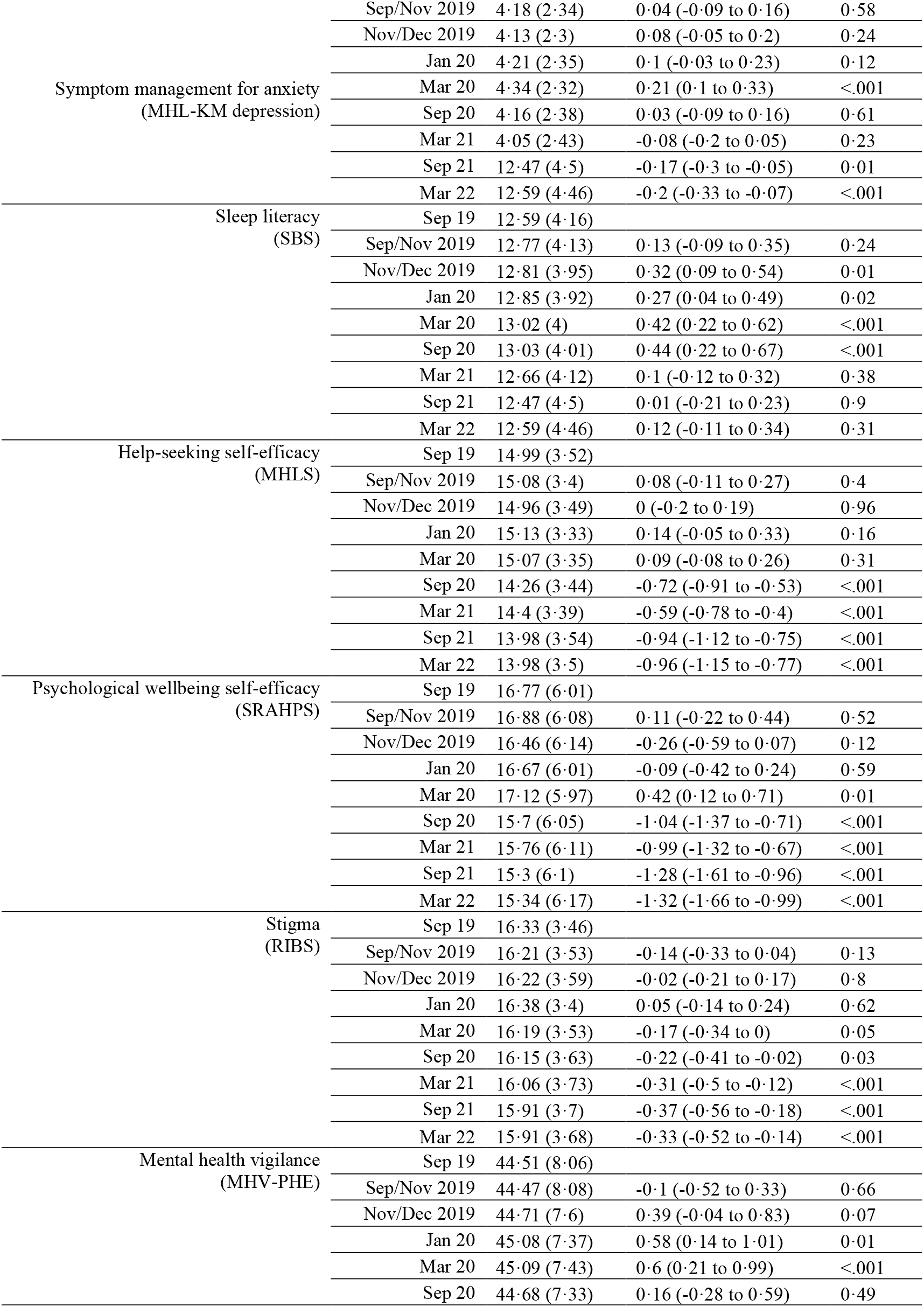
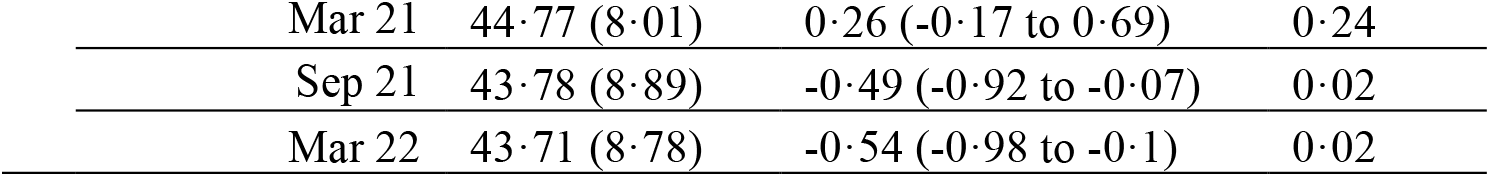
Changes in mental health literacy over time.

However, we observed reversal in these changes beyond March 2020. There was a small decline for symptom recognition for depression (MHL-REC depression) (d = −0·24; 95% CI −0·39 to −0·09; P = 0·002), symptom recognition for anxiety (MHL-REC anxiety) (d = −0·17; 95% CI −0·32 to −0·03; P = 0·021), and symptom management for depression (MHL-ACT depression) (d = −0·12; 95% CI −0·24 to 0·000003; P = 0·05) from Sep 2019 to Mar 2021, during the 6 months of the first pandemic lockdown in the UK. Symptom management for anxiety (MHL-ACT anxiety) showed no change from Sep 2019 scores by March 2020 (d = 0·03; 95% CI −0·09 to 0·16; P = 0·22), which suggests a decline in this score from Mar 2020. Mental health vigilance (MHL-VIG) showed a return to Sep 2019 levels for Sep 2020 (d = 0·15; 95% CI −0·27 to 0·57; P=0·49) and 8 (d = 0·25; 95% CI 0·16 to 0·67; P=0·24). Sleep literacy (SBS) also follows/ed this trend, wherein scores from Mar 2021 showed no difference compared to Sep 2019 scores (d = 0·08; 95% CI −0·13 to 0·30; P = 0·42). Help-seeking self-efficacy (MHLS) scores demonstrated moderate decline for Sep 2020 (d = −0·72; 95% CI −0·91 to −0·53; P<0·001) and Mar 2021 (d = −0·59; 95% CI −0·77 to −0·4; P = 0·001). Psychological wellbeing self-efficacy (SRAHPS) also exhibited a large decline to Sep 2020 (d = −1·03; 95% CI −1·37 to −0·7; P<0·001) and Mar 2021 (d = −0·99; 95% CI −1·32 to −0·66; P<0·001) compared to Sep 2019. Stigma related to mental health disorders (RIBS) showed a small decline from Sep 2019 to Mar 2020 (d = − 0·17; 95% CI −0·34 to 0; P = 0·046), Sep 2020 (d = −0·22; 95% CI −0·41 to −0·03; P = 0·025), and Mar 2021 (d = −0·31; 95% CI −0·5 to −0·12; P = 0·001).

By Mar 2022, all MHL outcomes except for sleep literacy (d = 0·12, 95% CI −0·11 to 0·34, P = 0·31) had shown a decline. There was strong evidence that symptom recognition for stress (d = −0·19, 95% CI −0·34 to −0·05, P = 0·01), depression (d = −0·3, 95% CI −0·45 to −0·14, P<0·001), and anxiety (d = −0·17, 95% CI −0·32 to −0·02, P = 0·03) has declined since September 2019. Similar declines are seen for symptom management for stress (d = −0·26, 95% CI −0·39 to −0·14, P<0·001), depression (d = − 0·31, 95% CI −0·44 to −0·19, P<0·001), and anxiety (d = −0·2, 95% CI −0·33 to −0·07, P<0·001). Help-seeking self-efficacy (d = −0·94, 95% CI −1·15 to −0·77, P<0·001) and psychological well-being self-efficacy outcomes have also shown to decline (d = −1·32, 95% CI −1·66 to −0·99, P<0·001). Finally, there was strong evidence that stigma scores declined (d = −0·33, 95% CI −0·52 to −0·14, P<0·001) and mental health vigilance was also lower (d = −0·54, 95% CI −0·98 to −0·1, P = 0·02) over 30 months of the campaign.

### Objective 2: Campaign awareness

The total number of participants included in the analysis was 13,178, and excludes participants from survey waves Sep 2019, Sep 2021, Mar 2022 as prompted awareness of the campaign was not measured in these waves (n = 7257). Demographic groups with relatively higher campaign awareness were the 18-24 age group, women, Black people, and people in the semi-/unskilled manual occupation group from their respective categories (Table 1).

Campaign awareness had varying associations with MHL outcomes (Table 3 and Figure 1-3). Those who encountered the campaign had slightly higher scores for symptom management of depression (MHL-ACT depression) (d = 0·13; 95% CI 0·05 to 0·21) and anxiety (MHL-ACT anxiety) (d = 0·18; 95% CI 0·06 to 0·29), and mental health vigilance (MHL-VIG) (d = 0·95; 95% CI 0·64 to 1·25) compared to those who have not encountered the campaign. Those who encountered the campaign showed moderately higher help-seeking self-efficacy (MHLS) (d = 0·41; 95% CI 0·15 to 0·67) (Figure S2) than those who have not encountered the campaign. Campaign awareness was associated with lower desire for social distance from people with mental illness (RIBS) (d = 0·35; 95% CI 0·19 to 0·51) (Figure S4).

**Table 3:** IPD meta-analysis of campaign awareness and mental health literacy outcome.

| Outcomes | Timepoint | Effect (95% CI) | Weight | t2 (95% CI) |
| --- | --- | --- | --- | --- |
| Symptom recognition of stress (MHL-KR stress) | Oct 2019 | 0.06 (-0.17 to 0.29) | 17.71 | 0 (0 to 0.0271) |
|  | Nov/Dec 2019 | 0.04 (-0.19 to 0.26) | 17.76 |  |
|  | Jan 2020 | -0.01 (-0.25 to 0.22) | 16.52 |  |
|  | Mar 2020 | 0.07 (-0.13 to 0.27) | 23.46 |  |
|  | Sep 2020 | -0.18 (-0.47 to 0.12) | 10.64 |  |
|  | Mar 2021 | -0.05 (-0.3 to 0.21) | 13.9 |  |
|  | Overall | -0.01 (-0.18 to 0.09) | 100 |  |
| Symptom recognition of depression (MHL-KR depression) | Oct 2019 | -0.06 (-0.3 to 0.18) | 17.59 | 0.018 (0 to 0.15) |
|  | Nov/Dec 2019 | -0.02 (-0.26 to 0.23) | 17.38 |  |
|  | Jan 2020 | -0.05 (-0.3 to 0.2) | 16.65 |  |
|  | Mar 2020 | 0.22 (0.02 to 0.43) | 19.66 |  |
|  | Sep 2020 | -0.38 (-0.69 to -0.06) | 13.1 |  |
|  | Mar 2021 | -0.09 (-0.36 to 0.18) | 15.62 |  |
|  | Overall | -0.04 (-0.24 to 0.15) | 100 |  |
| Symptom recognition of anxiety (MHL-KR anxiety) | Oct 2019 | 0.24 (0.01 to 0.48) | 17.19 | 0.02 (0 to 0.18) |
|  | Nov/Dec 2019 | 0.07 (-0.17 to 0.3) | 17.32 |  |
|  | Jan 2020 | 0.01 (-0.24 to 0.25) | 16.85 |  |
|  | Mar 2020 | 0.2 (0 to 0.4) | 19.37 |  |
|  | Sep 2020 | -0.37 (-0.67 to -0.06) | 13.63 |  |
|  | Mar 2021 | 0.08 (-0.19 to 0.34) | 15.64 |  |
|  | Overall | 0.06 (-0.16 to 0.27) | 100 |  |
| Symptom management of stress (MHL-KM stress) | Oct 2019 | 0.07 (-0.13 to 0.26) | 17.88 | 0.01 (0 to 0.09) |
|  | Nov/Dec 2019 | 0.07 (-0.13 to 0.27) | 17.38 |  |
|  | Jan 2020 | 0.18 (-0.03 to 0.39) | 16.69 |  |
|  | Mar 2020 | 0.12 (-0.05 to 0.3) | 20.33 |  |
|  | Sep 2020 | -0.28 (-0.54 to -0.03) | 12.5 |  |
|  | Mar 2021 | 0.12 (-0.1 to 0.34) | 15.21 |  |
|  | Overall | 0.06 (-0.09 to 0.22) | 100 |  |
| Symptom management of depression<br>(MHL-KM depression) | Oct 2019 | 0.14 (-0.05 to 0.33) | 18.57 | 0 (0 to 0.02) |
|  | Nov/Dec 2019 | 0.13 (-0.07 to 0.32) | 17.44 |  |
|  | Jan 2020 | 0.14 (-0.07 to 0.34) | 16.02 |  |
|  | Mar 2020 | 0.2 (0.03 to 0.37) | 23.08 |  |
|  | Sep 2020 | -0.05 (-0.3 to 0.21) | 10.53 |  |
|  | Mar 2021 | 0.13 (-0.09 to 0.35) | 14.35 |  |
|  | Overall | 0.13 (0.05 to 0.21) | 100 |  |
| Symptom management of anxiety<br>(MHL-KM anxiety) | Oct 2019 | 0.2 (0 to 0.39) | 18.24 | 0.02 (0 to 0.18) |
|  | Nov/Dec 2019 | 0.18 (-0.02 to 0.38) | 17.15 |  |
|  | Jan 2020 | 0.22 (0.01 to 0.43) | 15.98 |  |
|  | Mar 2020 | 0.22 (0.05 to 0.39) | 24.26 |  |
|  | Sep 2020 | -0.11 (-0.37 to 0.15) | 10.3 |  |
|  | Mar 2021 | 0.25 (0.02 to 0.47) | 14.07 |  |
|  | Overall | 0.18 (0.06 to 0.29) | 100 |  |
| Sleep literacy<br>(SBS) | Oct 2019 | 0.08 (-0.27 to 0.42) | 17.26 | 0.01 (0 to 0.09) |
|  | Nov/Dec 2019 | -0.18 (-0.52 to 0.17) | 17.46 |  |
|  | Jan 2020 | 0.1 (-0.25 to 0.44) | 17.15 |  |
|  | Mar 2020 | 0.34 (0.05 to 0.64) | 20.3 |  |
|  | Sep 2020 | -0.28 (-0.72 to 0.17) | 12.63 |  |
|  | Mar 2021 | -0.16 (-0.55 to 0.22) | 15.2 |  |
|  | Overall | 0.01 (-0.24 to 0.25) | 100 |  |
| Help-seeking self-efficacy<br>(MHLS) | Oct 2019 | 0.01 (-0.28 to 0.31) | 17.49 | 0 (0 to 0.02) |
|  | Nov/Dec 2019 | 0.25 (-0.06 to 0.56) | 16.74 |  |
|  | Jan 2020 | 0.59 (0.29 to 0.89) | 17.15 |  |
|  | Mar 2020 | 0.43 (0.18 to 0.69) | 19.18 |  |
|  | Sep 2020 | 0.72 (0.33 to 1.11) | 13.4 |  |
|  | Mar 2021 | 0.52 (0.19 to 0.84) | 16.03 |  |
|  | Overall | 0.41 (0.15 to 0.67) | 100 |  |
| Psychological wellbeing self-efficacy | Oct 2019 | -0.54 (-1.06 to -0.03) | 17.14 | 0 (0 to 0.04) |
| (SRAHPS) | Nov/Dec 2019 | -0.03 (-0.55 to 0.5) | 16.97 |  |
|  | Jan 2020 | 0.17 (-0.37 to 0.71) | 16.82 |  |
|  | Mar 2020 | 0.58 (0.13 to 1.03) | 18.15 |  |
|  | Sep 2020 | 1.19 (0.51 to 1.87) | 14.7 |  |
|  | Mar 2021 | 0.37 (-0.2 to 0.95) | 16.22 |  |
|  | Overall | 0.27 (-0.33 to 0.88) | 100 |  |
| Stigma related to mental disorders<br>(RIBS) | Oct 2019 | 0.37 (0.08 to 0.66) | 18.36 | 0.02 (0 to 0.23) |
|  | Nov/Dec 2019 | 0.32 (0.01 to 0.63) | 16.57 |  |
|  | Jan 2020 | 0.48 (0.18 to 0.78) | 17.4 |  |
|  | Mar 2020 | 0.47 (0.21 to 0.73) | 23.49 |  |
|  | Sep 2020 | 0.02 (-0.38 to 0.41) | 10.14 |  |
|  | Mar 2021 | 0.23 (-0.11 to 0.56) | 14.05 |  |
|  | Overall | 0.35 (0.19 to 0.51) | 100 |  |
| Mental health vigilance<br>(MHV-PHE) | Oct 2019 | 0.71 (0.02 to 1.39) | 16.09 | 0 (0 to 0.32) |
|  | Nov/Dec 2019 | 0.54 (-0.13 to 1.2) | 17.04 |  |
|  | Jan 2020 | 1.24 (0.57 to 1.9) | 17.3 |  |
|  | Mar 2020 | 0.92 (0.36 to 1.48) | 24.58 |  |
|  | Sep 2020 | 1.15 (0.34 to 1.96) | 11.52 |  |
|  | Mar 2021 | 1.26 (0.51 to 2.01) | 13.46 |  |
|  | Overall | 0.95 (0.64 to 1.25) | 100 |  |

**Figure 1:**
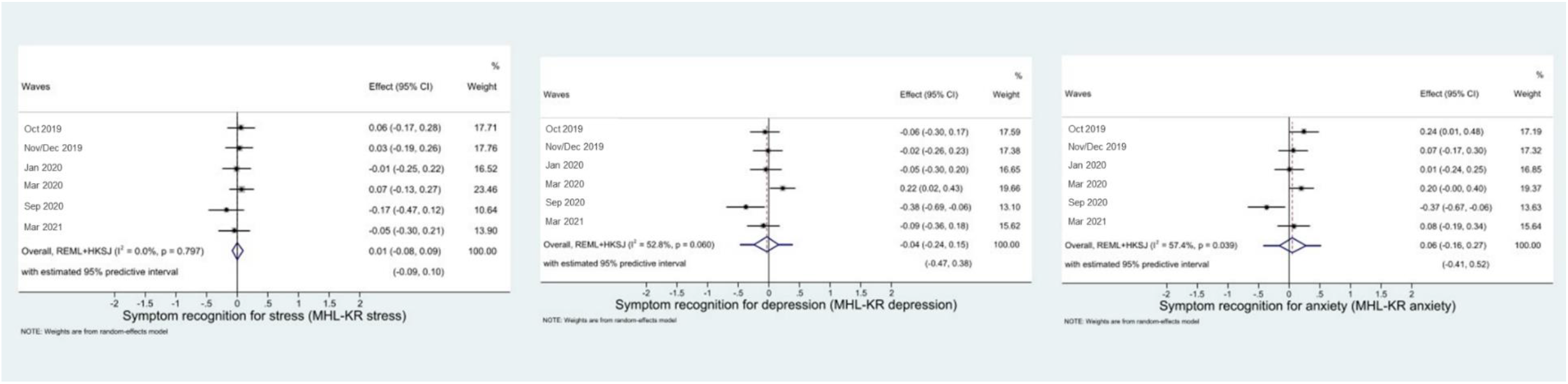
Effect size of awareness on symptom recognition of stress, depression, and anxiety (MHL-KR)

**Figure 2:**
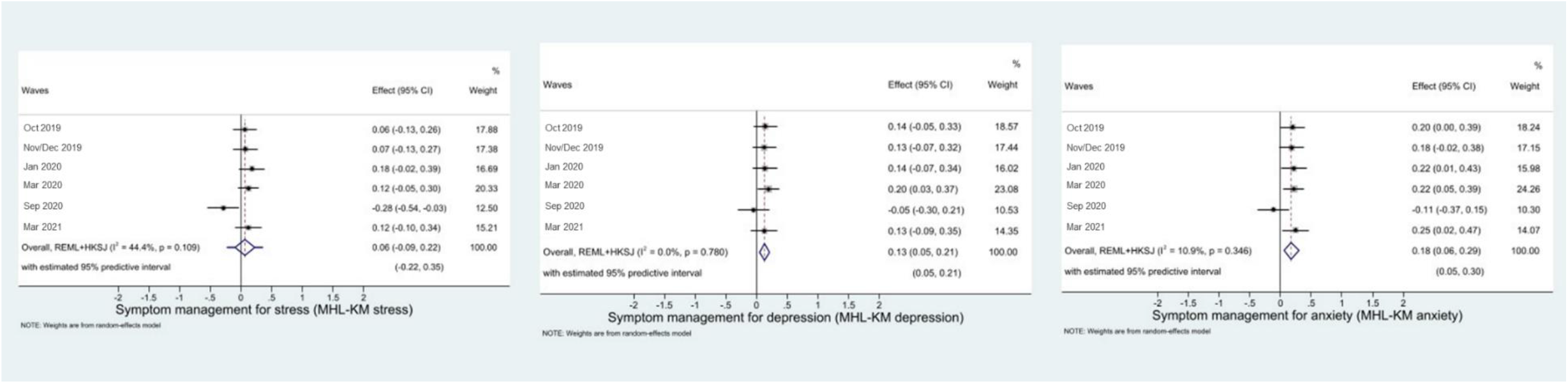
Effect size of awareness on symptom management of stress, depression, and anxiety (MHL-KM)

**Figure 3:**
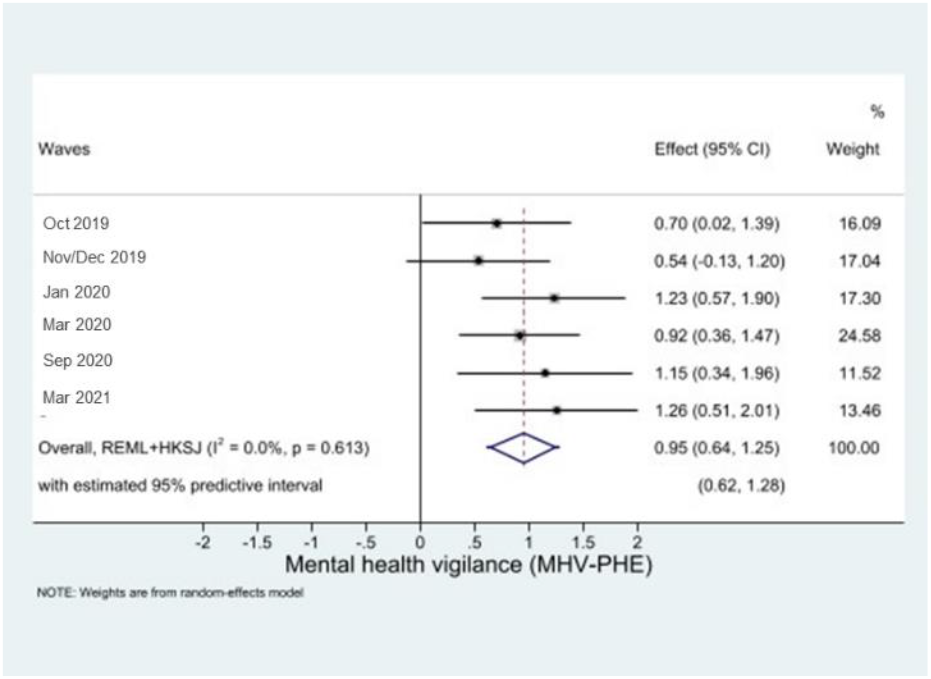
Effect size of awareness on mental health vigilance (MHV-PHE)

We observed little heterogeneity in associations between campaign awareness and symptom recognition of depression (MHL-REC depression) (τ^2^ = 0·02; 95% CI 0 to 0·15), anxiety (MHL-REC anxiety) (τ2 = 0·02; 95% CI 0 to 0·18), and stress (MHL-REC stress) (τ2 = 0·01; 95% CI 0 to 0·09), symptom management for anxiety (MHL-ACT anxiety) (τ2 = 0·02; 95% CI 0 to 0·18), sleep literacy (SBS) (τ2 = 0·01; 95% CI 0 to 0·02), and stigma (RIBS) (τ2 = 0·02; 95% CI 0 to 0·23). We did not observe any association between awareness and symptom recognition for stress, depression, anxiety, symptom management of stress, and help-seeking self-efficacy.

## Discussion

In terms of knowledge of symptoms and self-management, we saw small improvements over time in certain MHL outcomes such as symptom management for depression and anxiety, sleep literacy, and mental health vigilance during the first six months after EMM was launched, but this effect was not sustained 30 months after the launch of the campaign. Campaign awareness was positively associated with symptom management of depression, anxiety, and mental health vigilance. There were no observed relationships between campaign awareness and symptom recognition for stress, depression, anxiety, and symptom management of stress.

We observed small improvements for psychological wellbeing self-efficacy in the first six months of the campaign. Help-seeking self-efficacy levels remained similar throughout the first 18 months after campaign launch but deteriorated to be lower than pre-campaign levels after March 2020. Campaign awareness was not positively associated with psychological wellbeing self-efficacy but was associated with help-seeking self-efficacy.

Stigma in the form of desire for social distance showed no changes in the first six months of campaign activity. Thereafter we see increased desire for social distance. However, campaign awareness was positively associated with reduced stigma.

In accordance with the definition of MHL from Kutcher and colleagues, ^1^ these results indicate a short-lived improvement in knowledge for managing one’s own mental health problems, sleep-literacy, ability to promote one’s own mental health in the general population. We see better outcomes in knowledge of managing one’s own mental health, ability to seek help, and stigma related to mental health problems for those who are aware of the campaign.

### Strengths and limitations

We cannot establish temporality between the campaign and the outcomes. With campaign awareness, we do not know whether respondents benefitted from the campaign or whether people who were more mental health literate are more likely to remember the campaign.

While quota sampling ensures that subgroups in the population are represented within the study sample, it does not randomly draw samples from the population. ^21^ Therefore, the results may not be entirely generalisable to the adult population in England.

Finally, the population survey design does not measure the impact of use of the digital resources as opposed to the impact of the campaign at population level, as the subsample of people who visited the campaign website or interacted with digital resources on the website are also too small in proportion within the survey. Evaluation of the digital resource itself would require pre-post measurement of outcomes for resource users and a control group.

### Implications

Our data show an increase in MHL immediately before the COVID-19 pandemic and lower levels of MHL with the progression of the pandemic. One explanation is that experience of the pandemic normalised symptoms of common mental health problems due to difficult life circumstances and reduced self-efficacy due to limited access to certain activities for wellness or healthcare. As there was a decline in health literacy between pre- and post-pandemic in Japan, especially for those in lower economic groups, ^22^ this may suggest that difficult life circumstances may negatively impact general health literacy.

However, we should not expect a sudden and large effect size with public mental health campaigns over two and a half years. Consistent positive changes across stigma related to mental disorders were only seen five years after the launch of the Time to Change social marketing campaign. ^23^ The emphasis by PHE on helpful behaviours in relation to one’s own health mirrors this approach. The consistency of these insights suggests an early emphasis on behaviour should be considered for any population mental health campaign.

## Data Availability

All data produced in the present study are available upon reasonable request to the authors

## Contributors

Jane Sungmin Hahn: Formal analysis, Writing – original draft, Writing – review & editing. Kia Chong Chua: Methodology, Supervision, Writing – review & editing. Rebecca Jones: Methodology, Supervision. Claire Henderson: Conceptualisation, Methodology, Funding acquisition, Supervision, Writing − review & editing. The research in collaboration with NIHR Mental Health Policy Research Unit and PHE.

## Acknowledgements

We thank Public Health England for their collaboration; YouGov for collaboration on questionnaire development; and the grant holders for the NIHR Mental Health Policy Research Unit for their advice. We would also like to thank Dr Rebecca Jones, who sadly passed before the completion of this paper. Rebecca played a vital role in building the foundations of statistical analysis. We will miss her wise guidance and warm presence.

